# Estrogen interactions with breast cancer risk variants in regulatory DNA

**DOI:** 10.64898/2026.09.09.26362309

**Authors:** Ibtihal Elfaki, Laura N. Kellman, Robin M. Meyers, Luca Ducoli, Martina Fu, Kamal Obbad, Smarajit Mondal, Xue Yang, Douglas F. Porter, David L. Reynolds, Suhas Srinivasan, Taishi Nakase, Mineto Ota, Tania Fabo, Jordan M Meyers, Lu Yang, Nasa Sinnot-Armstrong, Kenneth E. Westerman, Linda Kachuri, Paul A. Khavari

## Abstract

Insight into population-level gene-environment (GxE) interactions is a major goal in understanding polygenic diseases, such as breast cancer. We integrated massively parallel reporter assays (MPRA), chromatin profiling, and network analysis to identify estrogen-responsive breast cancer variants. Screening 1,604 GWAS-identified breast cancer variants identified 73 estrogen-modulated SNVs (emSNVs). Chromatin accessibility modeling validated allele-specific effects, identifying variants that disrupt pioneer factor binding to chromatin and others that modulate transcription factor (TF) recruitment to pre-accessible enhancers. emSNV targets converged on pathways including mitochondrial metabolism, NF-κB signaling, and chromatin regulation. Aggregating emSNVs into a polygenic risk score (PRS_E2_) revealed interactions with reproductive risk factors in 13,026 post-menopausal BRCA cases and 108,265 controls, including age at first birth (p=0.0052); a control PRS lacking estrogen-responsive variants showed no interactions. This framework bridges molecular and epidemiological GxE studies to uncover variants whose disease associations depend on environmental context, with implications for understanding polygenic disease risk.

## INTRODUCTION

Breast cancer remains the most common cancer among women worldwide, with over 2 million new cases diagnosed annually^1^. While genetic susceptibility accounts for a substantial portion of disease risk, environmental and lifestyle factors, including reproductive history, hormone exposure, and metabolic factors, play critical roles in determining who develops disease. Understanding how genetic variants interact with these environmental exposures (**<u>GxE</u>**) is essential for identifying high-risk individuals, developing targeted prevention strategies, and explaining why some women with similar genetic profiles experience vastly different outcomes. However, despite their clinical importance, GxE interactions remain notoriously difficult to detect and characterize, limiting the ability to translate genetic discoveries into actionable risk predictions.

Genome-wide association studies (**<u>GWAS</u>**) have identified ∼ 300 loci associated with breast cancer risk.^2^ However, many standard analytical approaches assume genetic effects are constant across environmental contexts^3^. Environmental and hormonal factors such as estrogen exposure are established breast cancer risk factors, yet whether and how specific genetic variants interact with these exposures to modulate risk are still being defined^4,5^. Advances in functional genomics have improved the ability to characterize biological mechanisms underlying GWAS-identified associations. In tandem, computational tools and large-scale biobank datasets, which integrate genetic and phenotypic data, have provided unprecedented opportunities to explore how genetic susceptibility modifies the effects of environmental exposures on disease risk. Despite these advances, detecting GxE interactions in genome-wide interaction studies remains statistically challenging, often requiring large sample sizes to achieve adequate power.^6^ These statistical challenges motivate the need for complementary functional approaches to prioritize variants most likely to exhibit true GxE effects.

High throughput functional assays may offer an approach to address some of these limitations by identifying variants with context-specific regulatory effects in vitro. In this regard, Massively Parallel Reporter Assays (**<u>MPRA</u>**) can systematically screen noncoding variants identified by GWAS for functional, transcription-directing activity^7–13^ and thus provide a means of assessing the regulatory potential of SNVs with varying environmental stimuli^14^. Integrating MPRA findings with population data may offer novel insight into the molecular basis of GxE interactions, advancing understanding of how genetic and environmental factors may jointly influence breast cancer risk. Leveraging these tools enables the determination of how well GxE effects observed in vitro may recapitulate GxE interactions at the population level, an area that remains largely underexplored.

Here we use MPRA to identify noncoding BRCA risk-linked SNVs whose transcriptional activity is modulated by estrogen, a key hormonal risk factor for all BRCA subtypes^15^ and further use RNA-seq, ATAC-seq, and motif enrichment analysis to characterize the identified estrogen-modulated SNVs (emSNVs). Additionally, we explore chromatin accessibility, DNA looping, gene proximity, and eQTL data for these emSNVs to examine potential gene networks. Finally, we construct an emSNV-generated polygenic risk score (PRS_E2_) to further assess the GxE effects of these variants. Overall, we find that aspects of GxE risk may be modeled in 2D culture and point to the value of integrating functional genomics with epidemiological data in understanding polygenic disease risk.

## RESULTS

### Functional and epidemiological analyses of estrogen-modulated MPRA SNVs

To explore the effects of estrogen on the transcription-directing activity of breast cancer risk variants, we employed a multi-faceted approach integrating functional genomics with epidemiological analyses (**Fig. 1**). Genome-wide significant SNVs (P<5×10^-8^) were prioritized from the GWAS Catalog^16^ based on epigenomic features of regulatory DNA in human mammary cells (see **Methods**). The candidate list of variants was expanded to include SNVs in linkage disequilibrium (LD) (r^2^ > 0.8), yielding 1604 BRCA risk variants. After evaluating sample-level variation and library representation (**Supplementary Fig. 1a, Supplementary Fig. 1b**), we tested each SNV for synergistic transcription-directing effects with estrogen using MPRA **(Fig. 1a)**. To characterize the mechanisms underlying estrogen-modulated SNVs (emSNVs), we modeled TF binding and chromatin accessibility effects of these SNVs in silico (**Fig. 1b)** and performed RNA-seq and ATAC-seq in human MCF-7 breast cells under estrogen treatment to define the broader transcriptional and chromatin accessibility landscape in which emSNVs operate. Finally, we evaluated the epidemiologic relevance of emSNVs by assessing whether emSNV-based polygenic risk scores interact with estrogen-related risk factors to influence breast cancer risk in the UK Biobank (**Fig. 1c**).

**Figure 1:**
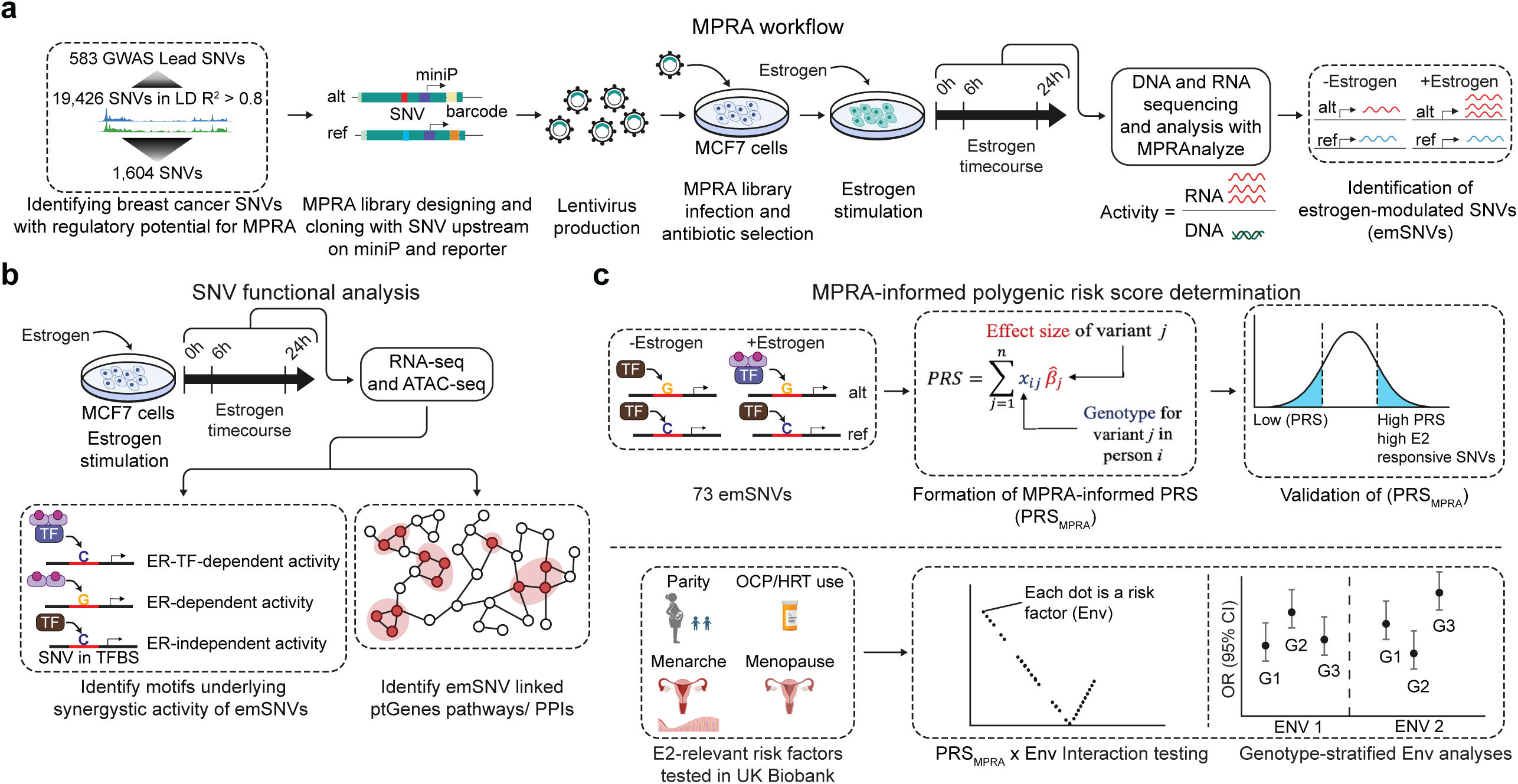
Estrogen-modulated transcriptional activity of breast cancer risk variants: functional characterization and assessment of genotype-estrogen interactions in biobank data. a) Massively parallel reporter assay (MPRA) workflow. Breast cancer (BRCA) single nucleotide variants (SNVs) were identified through GWAS studies and expanded in linkage disequilibrium (LD) to include SNVs in LD at r^2^ > 0.8 and then filtered through epigenomic peaks, although some SNVs were retained due to ancestry differences in GWAS and ENCODE data files. b) Functional characterization of motifs and genes associated with estrogen-modulated SNVs (emSNVs) using motif analysis, ATAC-seq, and RNA-seq. Estrogen (estrogen) influences transcriptional activity by altering TF (TF) binding, including estrogen receptor (ER), or by modifying chromatin accessibility. Motif analyses of emSNVs explore their potential impact on chromatin dynamics, while putative target genes are assigned to emSNVs and integrated with RNA-seq data to elucidate gene-regulatory networks. c) Generation of a polygenic risk score (PRS) derived from estrogen-sensitive SNVs (PRS_E2_) and assessment of its association with breast cancer risk.

### Estrogen modulates transcriptional activity of breast cancer risk variants

We identified 73 estrogen-modulated SNVs (emSNVs) showing statistically significant genotype-by-estrogen treatment interactions at FDR < 0.10 (42 at FDR < 0.05; **Fig. 2a, Extended Data 1**). Differences in ref vs alt allele activity were seen across conditions (**Supplementary Fig 1c**). emSNVs were broadly distributed across the genome (**Fig. 2b**), consistent with the widespread genomic effects of the estrogen receptor (ER) TF and diverse gene targets.^17^Among emSNVs, variants with the strongest GWAS effects on breast cancer susceptibility based on the Zhang et al. meta-analysis^18^ included rs9479091 (P=1.5×10^-37^), an intronic estrogen receptor 1 (*ESR1*) variant, and rs11836367 (P=6.0×10^-38^), which has been shown to contribute to breast cancer onset by modulating *NTN4* expression by increasing GATA3 binding.^19^

**Figure 2:**
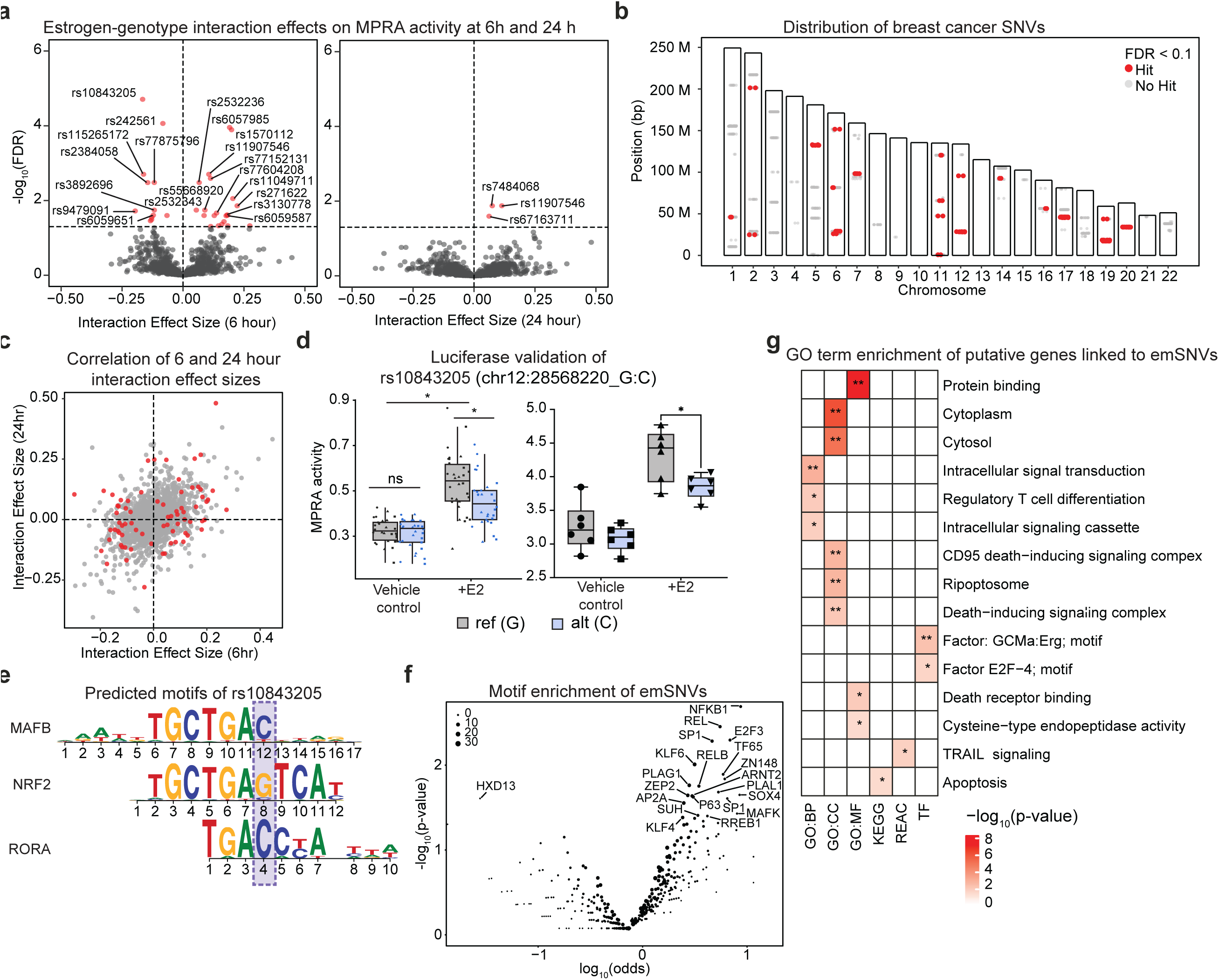
Breast cancer SNVs show genotype-by-estrogen interactions and enrich for apoptotic pathway regulation. a) Genotype-by-estrogen interaction effects on MPRA activity. Volcano plots showing interaction effect sizes (x-axis) versus statistical significance (y-axis, - log₁₀(FDR)) for all 1,604 tested variants at 6 hours (left) and 24 hours (right) post-estrogen treatment. Each point represents a variant; gray points indicate non-significant effects, while red points indicate significant genotype × estrogen interactions (FDR < 0.10, vertical dashed line at FDR = 0.10). Horizontal dashed lines mark FDR = 0.05. Positive effect sizes indicate increased allelic differences under estrogen treatment. Top variants are labeled (e.g., rs10843205, rs2453236, rs2384058 at 6h; rs7448061, rs11607546 at 24h). b) Genomic distribution of breast cancer SNVs. Histogram showing the chromosomal distribution of all tested breast cancer GWAS variants (n=1,604). Bars indicate position count per chromosome; red-filled bars highlight chromosomes containing emSNVs identified at FDR < 0.10, demonstrating broad genomic distribution consistent with widespread ER binding sites and diverse target genes. c) Correlation of interaction effects across timepoints. Scatter plot comparing genotype × estrogen interaction effect sizes at 6 hours (x-axis) versus 24 hours (y-axis). Red points indicate variants significant at either timepoint (FDR < 0.10); gray points are non-significant. Dashed lines mark zero effect. Moderate positive correlation (R = 0.44, p < 2.2×10⁻¹⁶) indicates partially overlapping but temporally dynamic regulatory effects, with most variants showing stronger interactions at 6 hours. d) Luciferase reporter assay of top emSNV rs10843205. Box plots showing normalized luciferase activity (MPRA activity) for reference (G, gray) and alternative (C, blue) alleles of rs10843205 (chr12:28568220_G/C) under vehicle control (left) or estrogen treatment (right) conditions. Under control conditions, alleles show minimal activity difference (ns, not significant). estrogen treatment reveals significant allele-specific enhancer activity, with the reference G allele driving higher transcriptional output than the C allele (p < 0.001). Individual data points represent biological replicates; boxes show median and interquartile range. e) Predicted TF motifs disrupted by rs10843205. Position weight matrices (PWMs) showing three TF binding motifs predicted to be altered by rs10843205: MAFB (top), NRF2 (middle), and RORA (bottom). Red box and arrows highlight the variant position within each motif. The reference G allele (position indicated) is predicted to favor NRF2 binding, while the alternative C allele favors MAFB and RORA binding. f) TF motif enrichment across all emSNVs. Volcano plot showing enrichment of TF binding motifs in emSNVs (FDR < 0.10, n=73) versus non-hit SNVs. X-axis shows log odds ratio (enrichment); y-axis shows -log₁₀(p-value). Each point represents a TF motif; labeled points indicate significantly enriched motifs including NF-κB 1, REL, SP1, E2F3, MAPK, KLF4/6, HXD13, PLAG1, and others. g) Gene Ontology enrichment of emSNV putative target genes. Heatmap showing significantly enriched GO biological process terms (rows) for genes linked to emSNVs through multiple orthogonal approaches (chromatin looping, eQTLs, proximity). Columns represent different gene-to-variant assignment methods: GWBP (genome-wide Bayesian fine-mapping), COJO (conditional and joint analysis), MAFB (MAF-based mapping), KEGGG (KEGG pathway mapping), REGC (regulatory circuit mapping), and TF (TF mapping). Color intensity indicates -log₁₀(p-value), with red representing strongest enrichment (p < 10⁻⁵). Asterisks denote significance levels (*p < 0.05, **p < 0.01). Strong enrichment is observed for protein binding, cytoplasmic signaling, intracellular signal transduction, apoptotic pathways (CD95 death-inducing signaling complex, ripoptosome, death receptor binding, TRAIL signaling), regulatory T cell differentiation, and TF activity (E2F4 and GCM1a:Erg motifs

Most variants showed significant estrogen-dependent allelic effects on regulatory activity at 6 hours (**Supplementary Fig 1d**), with a general pattern of attenuation at 24 hours. This temporal dynamic aligns with the known kinetics of ER-mediated transcriptional regulation, which predominantly occurs at earlier timepoints.^20^ There was a modest correlation between interaction effects across timepoints **(R=0.44, p<2.2×10⁻¹⁶; Fig. 2c)** indicating partially overlapping but temporally dynamic regulatory effects. We validated the top-ranked emSNV, rs10843205, using luciferase reporter assays. This variant is an eQTL for *CCDC91* in mammary tissue (GTEx), a gene previously associated with breast cancer susceptibility.^21^ Upon treatment with estrogen, the alternative C allele drove significantly higher transcriptional activity relative to the G allele (**Fig. 2d)**. In breast cancer GWAS, rs10843205-C was inversely associated with breast cancer risk (odds ratio (OR)=0.95, p=1.1×10^-11^). This suggests a mechanism where dampened estrogen-responsive enhancer activity at this locus potentially reduces *CCDC91* activity in response to estrogen and may be protective, consistent with a model where estrogen-driven upregulation of certain breast cancer susceptibility genes increases risk.

### emSNV motif analyses

To identify TFs that may mediate estrogen-dependent effects at emSNVs, we performed motif enrichment analysis using MotifbreakR.^22^ Estrogen is particularly relevant in this context as it can directly bind and facilitate ER DNA binding. In addition to ER DNA binding, ER has been shown to indirectly bind DNA through other TF associations^23–26^. We first examined a top-ranked variant, rs10843205, which is predicted to alter binding motifs for three TFs with established roles in estrogen signaling: NRF2, MAFB, and RORA (**Fig. 2e**). The reference G allele preferentially supports NRF2 binding, while the alternative C allele favors MAFB and RORA binding. NRF2, a master regulator of oxidative stress responses, is modulated by estrogen in ER-positive breast cancer cells, ^27–31^ and its pharmacological inhibition attenuates estrogen-mediated transcriptional effects.^30^ RORA, an orphan nuclear receptor with putative tumor suppressor function, has been associated with breast cancer, ^32–34^ though its mechanistic role in estrogen signaling remains incompletely defined. MAFB and related MAF family members interact with ER and have been implicated in breast cancer progression^35^. Chromatin immunoprecipitation footprinting data (ENCSR620CKC) further support binding of MAFK, a related MAF protein, at rs10843205, with preferential occupancy at the reference G allele (**Supplementary Fig. 1e**). These findings identify rs10843205 as an estrogen-responsive regulatory variant where allele-specific TF binding modulates enhancer activity in a hormone-dependent manner. Extending this analysis genome-wide, we examined TF motif enrichments across all 73 emSNVs relative to non-functional SNVs (**Fig. 2f, Extended Data 2**). emSNVs were significantly enriched for binding motifs of TFs with established roles in estrogen signaling, including NFKB/REL family members. SP1, E2F, MAF, and KLF4/KLF6. NFKB/REL family members exhibit well-characterized bidirectional crosstalk with ER signaling in breast cancer. ^36–38^ SP1 is essential for ER-driven transcriptional responses, ^39–42^ while E2F TFs regulate cell cycle progression and serve as markers of breast cancer aggressiveness.^43^ KLF4 similarly modulates estrogen-dependent gene regulation.^44^ Condition-specific motif enrichments showed similar patterns (**Supplementary Fig. 1f**). These motif enrichments nominate candidate TFs for future functional investigation at emSNV loci and suggest that estrogen-dependent gene regulation is shaped by cooperative or competitive interactions among multiple regulatory factors beyond ER itself.

### emSNV-modulated ptGene network

To identify biological processes and potential therapeutic targets associated with emSNVs, we constructed a protein-protein interaction network of putative target genes (ptGenes) assigned to emSNVs through multiple orthogonal approaches (see Methods, **Extended Data 3**). Genes linked to breast cancer-associated SNVs that could be mapped to STRING were pruned to remove isolated nodes, resulting in a network (**Supplementary Fig. 2**) of 267 proteins (nodes) and 1052 interactions (edges). There are 38 proteins that interact with anti-neoplastic drugs, 17 TFs, and 78 SNV-linked proteins that are GxE hits. Network analysis identified several high-degree nodes that act as hubs, e.g., H4C1, H4C6 and H4C11 among similar highly connected nodes. This suggests that the identified hub proteins may modulate multiple biological processes and that changes to their function can affect many interaction partners.^45^ Notably, H4C1, a central hub in the network, has been reported as a biomarker of neoadjuvant response and prognosis in breast cancer^46^ Consistent with the enrichment of histone hubs in the network, previous studies have shown that estrogenic compounds produce similar proteomic changes selectively in ERα-positive MCF-7 cells, with histones H2A, H2B, H3 and H4 among the most strongly responsive proteins, whereas ER-negative cell lines show neither histone induction nor a proliferative response.^47^ This concordance supports the idea that estrogen-responsive emSNVs converge on chromatin-based regulatory mechanisms in breast cancer. Edge betweenness quantifies interactions that serve as links to various neighborhoods of the interaction network.^48^ Bridging paths such as UBA52-RPS23 and H3C12-HSPA4, have large edge betweenness values and a functional change in one of these genes could have a cascading effect in different parts of the network. Some of the modules identified by Markov clustering were functionally enriched with GO: Biological Process, seven of which are highlighted (**Supplementary Fig. 2**).

Gene co-expression largely mirrored patterns in modules including nucleosome assembly, mitochondrial translation, oxidative phosphorylation, NF-κB signaling, mitotic checkpoint signaling, and T-cell reporter signaling. Nucleosome assembly plays a critical role in estrogen-mediated gene regulation in breast cancer.^49,50^ Mitochondrial translation is increasingly recognized as important in breast cancer metabolism and estrogen response.^51,52^ Oxidative phosphorylation is a key metabolic vulnerability in ER-positive breast cancer, particularly in endocrine- and CDK4/6-resistant tumors, where its upregulation is linked to estrogen signaling, poor prognosis, and sensitivity to targeted inhibition.^53^ As previously discussed, NF-κB signaling interacts significantly with estrogen signaling in breast cancer and its constitutive activation in advanced or hormone-independent tumors promotes progression, metastasis, and immune evasion.^38,54^ Estrogen receptor signaling suppresses T-cell infiltration in breast cancer by dampening pro-inflammatory pathways, yet patients with high ER activity respond well to endocrine therapy, suggesting that reversing this immunosuppression could improve immunotherapy outcomes.^54^ Further, ongoing research also suggests ERα directly regulates mitotic checkpoint genes to maintain chromosomal stability, but dysregulation of this pathway is linked to poor prognosis in hormone receptor-positive breast cancer.^55^ A number of ptGenes are targets of approved medications,^56^ providing a rationale to support their future assessment in experimental models of estrogen-driven breast neoplasia.

### Estrogen-responsive transcriptional programs and emSNV target gene functions

To integrate estrogen-regulated transcriptional responses with estrogen-modulated variant activity, we performed RNA-seq on MCF7 cells treated with estrogen or vehicle control at 6 and 24 hours **(Supplementary Fig. 3a–b; Extended Data Fig. 4**). Unsupervised clustering revealed distinct patterns of gene induction and repression at 6 and 24 hours (**Supplementary Fig. 3c**), and an UpSet plot highlighted the overlap among these temporal expression programs (**Supplementary Fig. 3d**). Estrogen altered the expression of 1,717 mRNAs (|log₂FC| > 0.5, adjusted P < 0.05), with enriched transcripts including canonical estrogen-responsive genes such as GREB1, TFF1, PGR, PDZK1 (**Extended Data Fig. 4**) involved in cell-cycle regulation and hormone signaling (**Supplementary Fig. 3e-f; Supplementary Fig. 4a-c**).

To assess whether emSNVs influence estrogen-responsive transcription, we examined overlap between emSNV-linked ptGenes and estrogen-regulated differentially expressed genes (DEGs). Of 200 emSNV-linked ptGenes, 50 (25%) were also DEGs (**Supplementary Fig. 3f**). This partial overlap is consistent with MPRA measuring direct cis-regulatory activity of isolated elements, while endogenous gene expression reflects integration of multiple regulatory inputs including distal enhancers^57,58^, chromatin context,^59–61^ and post-transcriptional mechanisms.^62,63^

### Functional insights into emSNV-putative target genes

To gain functional insight into the biological pathways influenced by emSNVs, we performed gene ontology (GO) enrichment analysis on emSNV ptGenes. This analysis revealed strong enrichment in cytoplasmic signaling networks and protein binding, with a prominent cluster of pathways implicated in programmed cell death (**Fig. 2g**). In particular, components of the TRAIL signaling pathway, CD95 death-inducing signaling complex (DISC), cysteine-type endopeptidase activity, and ripoptosome were significantly overrepresented, suggesting these estrogen-interacting variants may influence key regulators of apoptosis. Each of these apoptotic pathways are also tightly linked to NF-κB signaling. TRAIL and Fas (CD95) are members of the death receptor subset of TNF receptors and can promote apoptosis under certain conditions.^64^ Similarly, the ripoptosome, comprising RIPK1, FADD, and caspase-8, can mediate both cell death and NF-κB activation in a context-dependent manner.^65^ We also saw enrichment for E2F-related regulators (e.g., *E2F4 TF complex*). E2F proteins govern cell cycle progression and apoptosis and are known to be modulated by ER signaling^66^ Enrichment was also observed for TF motifs associated with ERG, which is repressed by ER alpha and inhibits ER-dependent transcription.^67^ This is consistent with the emSNV motif analysis which revealed significant enrichment of binding site alterations for NF-κB family TFs (NF-κB 1, RELA, RELB) within emSNVs and E2F factors (**Fig. 2f**). These results suggest that emSNVs may alter estrogen-dependent transcriptional control of proliferation and death.

### Estrogen-dependent chromatin accessibility landscapes enrich for breast cancer heritability

To understand how emSNVs influence gene regulation through chromatin-mediated mechanisms, we generated ATAC-seq profiles from MCF-7 cells treated with vehicle control, 6-hour estrogen, or 24-hour estrogen treatment. All samples demonstrated robust chromatin accessibility signal quality (**Supplementary Fig. 5**). To assess whether these estrogen-responsive chromatin states reflect disease-relevant regulatory activity, we applied stratified LD score regression (S-LDSC) to partition breast cancer heritability according to condition-specific ATAC-seq peak sets. All three conditions showed statistically significant heritability enrichment for overall breast cancer, with 6-hour estrogen treatment exhibiting the strongest signal (enrichment p = 3×10⁻⁴), while no enrichment was observed for peaks from the unrelated K562 cell line (**Fig. 3a, Supplementary Fig. 6**). Similar enrichment patterns were observed across breast cancer subtypes, including ER+, ER-, Luminal A, and triple-negative disease (Supplementary Fig. 6, Extended Data 5). These findings establish that estrogen-induced chromatin remodeling in MCF-7 cells captures regulatory elements relevant to breast cancer susceptibility, with peak regulatory activity occurring at 6 hours post-treatment.

**Figure 3:**
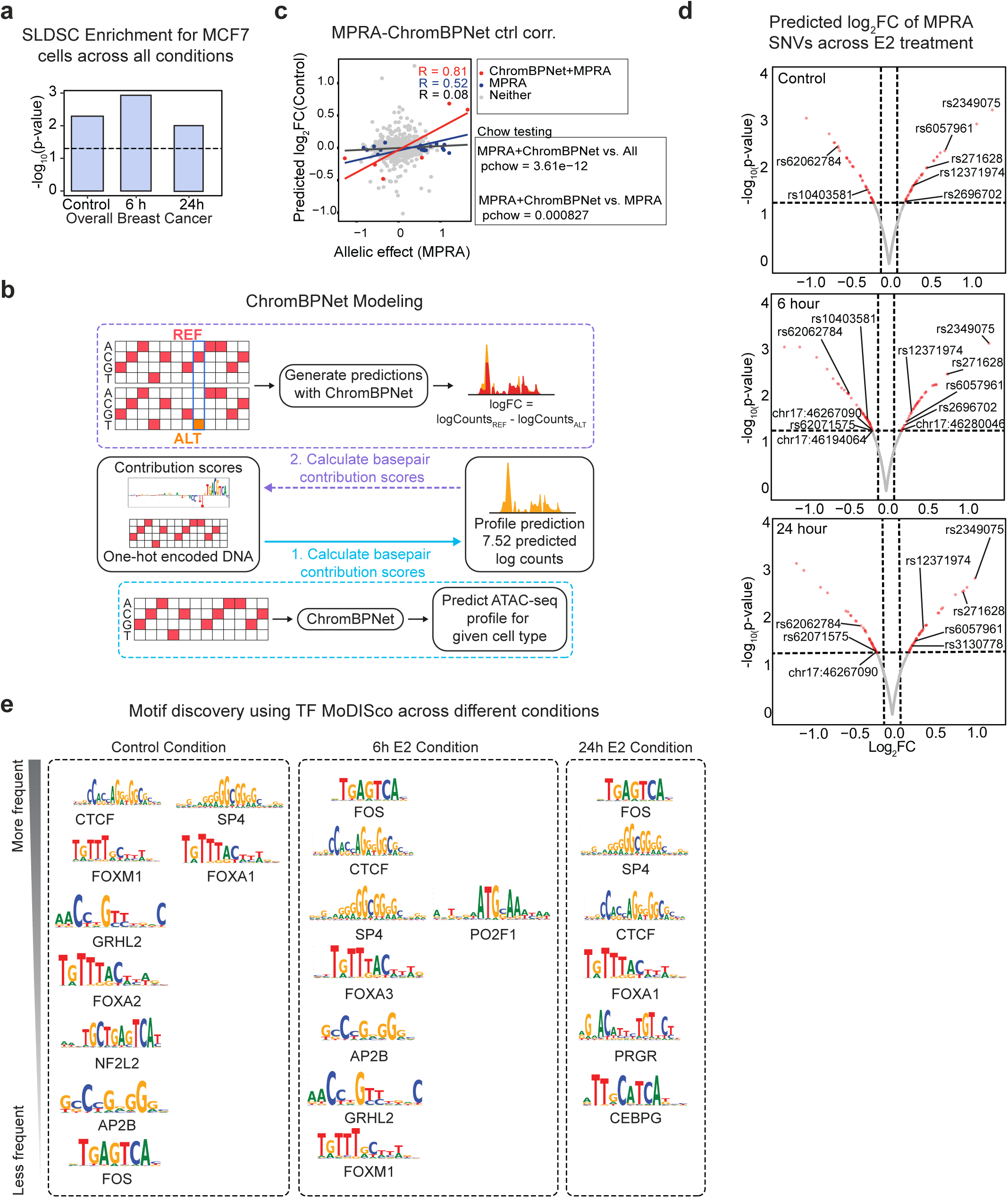
Chromatin accessibility modeling reveals estrogen-dependent regulatory mechanisms and temporal TF dynamics. a) ChromBPNet workflow. DNA sequence and chromatin accessibility (ATAC-seq) data are used to train deep learning models that predict allele-specific chromatin effects. One-hot encoded DNA sequences generate predicted accessibility profiles and importance scores, which are used to calculate basepair contribution scores and predict variant effects on chromatin state. b) Breast cancer heritability enrichment in estrogen-responsive chromatin. Stratified LD score regression (S-LDSC) shows significant enrichment of overall breast cancer heritability in ATAC-seq peaks from vehicle control, 6-hour estrogen, and 24-hour estrogen conditions, with strongest signal at 6 hours (p=3×10⁻⁴), validating disease relevance of estrogen-responsive chromatin states. c) ChromBPNet predictions correlate with MPRA effects. Scatter plot comparing predicted chromatin accessibility effects (ChromBPNet, y-axis) with experimentally measured allelic effects (MPRA, x-axis) in vehicle control condition. Gray points show all variants; red points indicate variants significant in both assays (FDR < 0.10). Overall correlation is modest (R=0.08) but variants significant in both assays show strong concordance (R=0.81, Chow test p=3.6×10⁻¹²), demonstrating ChromBPNet captures biologically meaningful allelic effects on chromatin accessibility. d) Predicted chromatin effects of MPRA-significant SNVs across estrogen treatment. Volcano plots showing ChromBPNet-predicted log fold-change in accessibility (x-axis) versus MPRA significance (y-axis, -log₁₀ p-value) for emSNVs across control (top), 6-hour (middle), and 24-hour (bottom) estrogen conditions. Vertical dashed lines mark log₂FC thresholds; horizontal dashed lines mark p=0.05. Several variants show condition-specific predicted accessibility changes (labeled), including rs2349075, rs6057961, and rs2071628, demonstrating that chromatin modeling identifies estrogen-dependent regulatory mechanisms in native genomic contexts. (e) Temporal TF programs revealed by TF-MoDISco. Position weight matrices of TF motifs identified by TF-MoDISco analysis of ChromBPNet importance scores, grouped by frequency (most to least frequent, top to bottom) across vehicle control (left), 6h estrogen (center), and 24h estrogen (right) conditions. Pioneer factors (CTCF, FOXA family) and architectural proteins (SP4) appear across all conditions. Condition-specific factors include NF2L2 (control only, chromatin priming), POU2F1 (6h, ER co-activator), and PR/GR and CEBPG (24h, sustained hormone response), revealing temporally ordered waves of TF engagement driving estrogen-responsive chromatin remodeling.

### Allele-specific chromatin accessibility reveals complementary regulatory mechanisms

We next applied ChromBPNet,^68^ a deep learning framework that models sequence determinants of chromatin accessibility, to predict allele-specific effects of emSNVs within endogenous genomic contexts (**Fig. 3b**). ChromBPNet was trained separately on ATAC-seq data from each condition (vehicle, 6h estrogen, 24h estrogen), enabling condition-specific predictions of how variants alter chromatin state. To benchmark these predictions, we compared ChromBPNet variant effect scores with experimentally measured MPRA allelic effects. Genome-wide correlation was modest (R = 0.08), while variants significant in MPRA showed minimal correlation (R = 0.52), and variant significant in both assays showed strong concordance (R = 0.81, Chow test p = 3.6×10⁻¹², **Fig. 3c, Extended Data 1, 6**), consistent with previously reported values^68,69^. This may reflect fundamental differences in these analyses: MPRA measures transcriptional output from integrated reporters, while ChromBPNet predicts chromatin accessibility at native loci. Variants can influence transcriptional activation without altering chromatin accessibility, and vice versa because these regulatory layers are mechanistically separable. Variants disrupting pioneer factor motifs (e.g., FOXA1) may substantially alter chromatin opening but show modest MPRA effects if the pioneer factor itself has weak transactivation activity. Conversely, variants affecting strong transcriptional activators recruited to pre-accessible chromatin (e.g., NF-κB) may exhibit robust MPRA signals without influencing chromatin accessibility. The strong concordance observed specifically among variants significant in both assays indicates that a subset of emSNVs simultaneously influence both regulatory layers, likely representing composite regulatory elements where chromatin accessibility and transcriptional activation are tightly coupled. This integration permitted distinction between accessibility-modulating variants (ChromBPNet-positive) and recruitment-modulating variants (MPRA-positive, ChromBPNet-neutral) and dual-mechanism variants (significant in both assays). Several emSNVs showed condition-specific predicted accessibility changes, including rs2349075, where the alternative allele was predicted to increase chromatin accessibility specifically upon estrogen treatment (**Fig. 3d, Extended Data 6-8**). These results demonstrate that ChromBPNet captures biologically meaningful allelic effects on chromatin state and, when integrated with MPRA, provides a multi-layered view of how variants modulate estrogen-responsive gene regulation.

### Temporal dynamics of TF programs driving estrogen-responsive chromatin remodeling

To identify which TFs mediate estrogen-dependent chromatin accessibility changes at emSNV loci, we applied TF-MoDISco, an unbiased motif discovery algorithm, to ChromBPNet importance scores (**Fig. 3e**). This analysis revealed distinct temporal programs of TF engagement across conditions. Pioneer factors and architectural proteins were detected across all timepoints, including FOXA family members (FOXA1/2/3), CTCF, and SP/KLF family factors (SP4), consistent with their roles in establishing and maintaining chromatin accessibility at enhancers. Condition-specific factors emerged at each timepoint, reflecting temporally ordered transcriptional responses. In vehicle-treated cells, NF2L2 (NRF2) motifs were prominent, consistent with its role as a chromatin priming factor at ERα-bound super-enhancers enhancers.^70^ At 6 hours post-estrogen, POU2F1 (OCT1) motifs appeared, matching its known function in inducing ESR1 transcription.^71^ By 24 hours, progesterone/glucocorticoid receptor (PR/GR) half-sites and CEBPG motifs emerged. PR is a canonical ERα interacting partner,^72^ while the related factor CEBPD has been shown to interact with ER signaling.^73^ These temporal dynamics suggest that estrogen triggers sequential waves of TF recruitment, beginning with chromatin priming factors, followed by early ER-interacting TFs, and culminating in sustained activity of hormone receptor networks. This ordered activation likely reflects the hierarchical nature of enhancer activation in estrogen-responsive tissues.

### Distinct regulatory modes: chromatin accessibility vs. TF recruitment

To integrate these findings with the earlier motif analysis (**Fig. 2f**), we compared TFs identified by sequence-based motif prediction (motifbreakR) versus accessibility-based inference (TF-MoDISco). Notably, several TF families enriched in motifbreakR analysis, including NF-κB/REL and E2F3, were absent from TF-MoDISco results. This discordance reflects distinct regulatory modes of action. Pioneer factors such as FOXA and AP-1 family members, recovered by both methods, actively remodel chromatin and drive accessibility changes.^74,75^ In contrast, recruited factors like NF-κB typically bind to pre-accessible enhancers established by pioneer factors^76^ and may not themselves drive measurable accessibility changes despite regulating transcription.^77^ This distinction has important mechanistic implications: emSNVs that disrupt pioneer factor motifs (detected by both methods) likely alter chromatin accessibility and affect all subsequently recruited TFs at that locus. Conversely, variants disrupting recruited factor motifs (motifbreakR-only) may modulate transcription without changing accessibility profiles. Together, these analyses indicate that estrogen-responsive variants influence gene regulation through at least two mechanisms: (1) accessibility-dependent effects mediated by altered pioneer factor binding, and (2) accessibility-independent effects via differential recruitment of secondary TFs to pre-established regulatory elements. This layered regulatory logic may explain why only a subset of emSNVs show concordant effects across MPRA, ChromBPNet, and RNA-seq analyses.

### MPRA-informed Polygenic Risk Score reveals GxE at the population level

To examine whether the interactions identified using MPRA generalize at the population level, independent emSNVs (LD r2<0.10) were combined into a polygenic risk score (PRS_E2_) for interaction testing in 13,026 post-menopausal BRCA cases and 108,265 controls of European ancestry from the UK Biobank (**Fig. 4a**). A control PRS was also developed that included matched genome-wide significant SNVs that did not exhibit statistically significant responses to estrogen stimulation (PRS_E2-null_; see Methods). Interaction analyses focused on established estrogen-related and reproductive risk factors that were at least nominally associated with breast cancer in the UK Biobank, including parity (≥1 live births: OR=0.85, 95% confidence interval (CI): 0.81-0.89), age at first birth (≥30 years: OR=1.13, 95% CI: 1.10-1.21), age at menarche (>13 years: OR=0.94, 95% CI: 0.90-0.97), age at menopause (OR = 0.99, 95% CI = 0.99-0.99), and hormone replacement therapy (HRT) use (OR=0.87, 95% CI: 0.84-0.90). (**Extended Data 9**).

**Figure 4:**
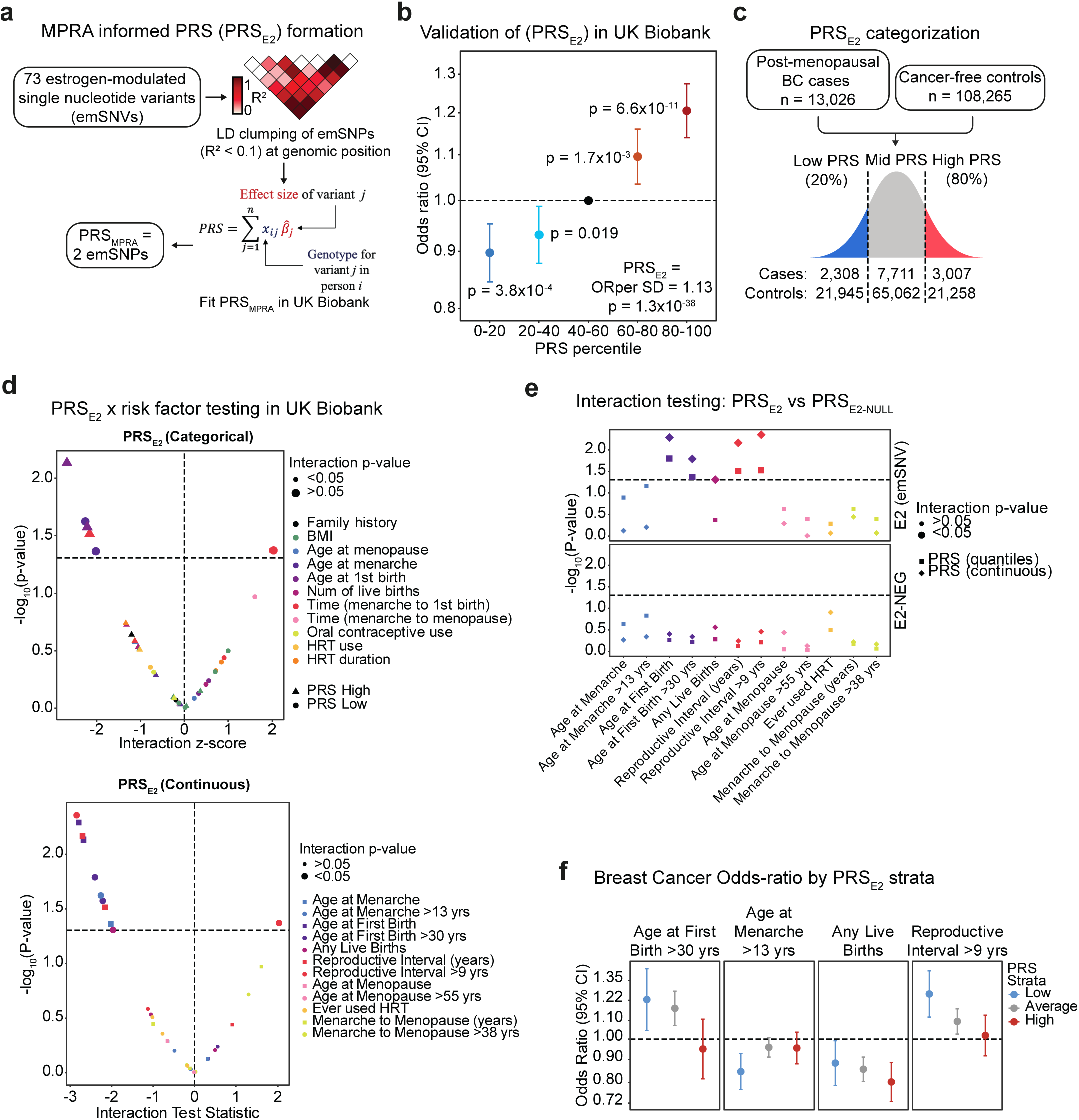
MPRA-informed polygenic risk score reveals gene-environment interactions in UK Biobank. a) PRS_E2_ construction workflow. 73 estrogen-modulated SNVs (emSNVs) identified at FDR < 0.10 were pruned for linkage disequilibrium (r² < 0.10), yielding 22 independent variants. Each variant was weighted by its effect size (β) from the Zhang et al. breast cancer GWAS meta-analysis to generate PRS_E2_. A matched negative control PRS (PRS_E2-NULL_) comprising 22 breast cancer risk variants from the same loci without estrogen-responsive activity was constructed similarly. b) PRS_E2_ validation in UK Biobank. Women in the top quintile (>80%) have elevated risk compared to average (40-60%, reference dashed line. c) PRS_E2_ stratification strategy. Distribution of PRS_E2_ in cases (red, n=13,026) and cancer-free controls (gray, n=108,265) showing shift toward higher PRS values in cases. Analysis groups: low PRS (≤20th percentile, blue), mid PRS (20-80th percentile, gray reference), high PRS (>80th percentile, red), with sample sizes shown. d) PRS × risk factor interaction testing. Interaction p-values (-log₁₀ scale) for PRS_E2_ with reproductive and hormonal risk factors, modeled categorically (top) and continuously (bottom). X-axis shows interaction test statistic (z-score); colors indicate high (red) vs. low (blue) PRS strata. Horizontal dashed line marks p=0.05; vertical dashed line marks z=0. Significant interactions (p < 0.05) observed for age at first birth (categorical: p=0.016; continuous: p=5.2×10⁻³), reproductive interval (categorical: p=0.031; continuous: p=6.9×10⁻³), and parity (categorical: p=0.049). All interaction terms are negative, indicating attenuation of risk factor effects in high PRS_E2_ individuals. e) Comparison of PRS_E2_ vs. PRS_E2-NULL_ interactions. Interaction p-values for PRS (top, PRS_E2_) and PRS_E2-NULL_ across reproductive risk factors, modeled categorically (diamonds) and continuously (circles). Colors represent PRS modeling approach. PRS_E2_ shows multiple significant interactions (above dashed p=0.05 line) with age at first birth, reproductive interval, and related factors. PRS_E2-NULL_ shows no significant interactions with any risk factor, demonstrating that gene-environment interactions are specific to functionally estrogen-responsive variants, not general breast cancer risk variants. f) Stratified risk factor associations by PRS_E2_. Odds ratios (95% CI) for reproductive risk factors within PRS_E2_ strata: low (blue, ≤20%), average (gray, 20-80% reference), high (red, >80%). Late age at first birth (>30 yrs) increases risk in low PRS (OR=1.22, p=0.012) and average PRS (OR=1.17, p=4.4×10⁻⁴) but not high PRS (OR=0.95, p=0.51). Long reproductive interval (>9 yrs) similarly shows risk effects in low PRS (OR=1.26, p=1.3×10⁻⁴) but not high PRS (OR=1.02, p=0.73).

Variants in each PRS were weighted by their marginal effects on breast cancer risk obtained from a GWAS meta-analysis of 133,384 cases and 113,789 controls by Zhang et al.^18^ The MPRA activity, breast cancer risk association, and associated gene(s) for each SNV considered for inclusion in the PRS are summarized in Table 1. Of the 24 emSNVs included in PRS_E2_, most had effects of similar magnitude and direction for overall breast cancer and specific subtypes, such as luminal A-like, luminal B/HER2-negative-like, luminal B-like, HER2-enriched-like and triple-negative or basal-like (**Extended Data 10**). Heterogeneous effects were observed for rs7118391-T, which was inversely associated with HER2 enriched-like and luminal B/HER2-negative-like subtypes, but positively associated with the remaining subtypes. Despite including a relatively low number of variants, both PRS_E2_ (OR= 1.13, p = 1.3×10^-38^) and the emSNV-negative control PRS_E2-NEG_ (OR per SD = 1.12, p = 8.6×10^-32^) were significantly associated with post-menopausal breast cancer in UKB (**Extended Data 11**). Women in the top quantile (>80%) of the PRS_E2_ distribution had significantly higher breast cancer risk (OR=1.21, p=6.6×10^-11^) compared to those with an average burden of emSNVs in the 40^th^-60^th^ percentiles (**Fig. 4b**).

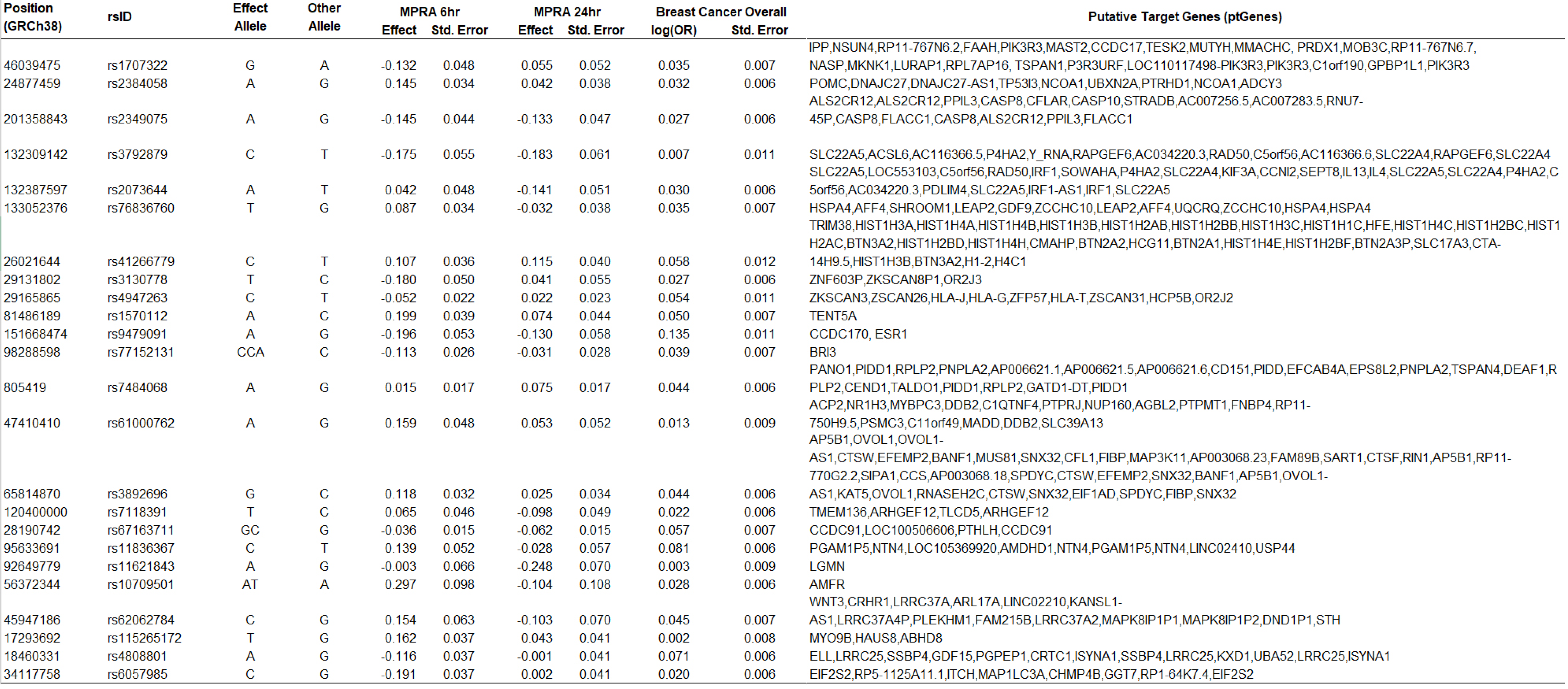

Analyses of PRS_E2_ were performed using both continuous and categorical specifications. For the categorical approach, PRS_E2_ was stratified into percentile-based groups (low: ≤20%, average: >20-80%, high: >80%) (**Fig. 4c**). Using both representations allowed capture of the full linear risk gradient while also enabling clinically interpretable group comparisons and detection of potential non-linear or threshold effects across the PRS distribution. When modeled as continuous predictors, statistically significant interactions with PRS_E2_ were observed for reproductive risk factors, such as having any live births (P_PRSxE_ =0.049), age at first birth (P_PRSxE_ =5.2×10^-3^), and reproductive interval (P_PRSxE_=6.9×10^-3^), but not factors related to menopause (**Fig. 4d; Extended Data 12**). All analyses produced negative interaction terms, suggesting that PRS_E2_ has an attenuating impact on the effect of each risk factor. Modeling continuous reproductive risk factors as categorical, to account for potential threshold effects or scale dependence, did not change the observed results (age at first birth >30 years: P_PRSxE_ =0.016; reproductive interval >9 years: P_PRSxE_ =4.4×10^-3^). When examining interactions across percentiles of PRS_E2_ (low: ≤20%, average: >20-80%, high: >80%), statistically significant interactions were observed for age at first birth (P_PRSxE_ = 0.016), reproductive interval (P_PRSxE_ = 0.031), and dichotomized versions of the same risk factors **(Fig. 4d; Extended Data 13**). Notably, no statistically significant interactions were observed for PRS_E2-NEG_ with any reproductive or menopause-related risk factors when PRS_E2-NEG_ was modeled as a continuous **(Extended Data 14)** or discrete variable **(Extended Data 15) (Fig. 4e)**.

To contextualize the observed interaction pattern, associations with breast cancer for relevant risk factors were estimated within strata of PRS_E2_ (low: ≤20%, average: >20-80%, high: >80%). Older age at first birth (≥30 years) was associated with increased breast cancer risk among women with low PRS_E2_ (OR=1.22, p=0.012) and average PRS_E2_ (OR=1.17, p=4.4×10^-4^), but not among women in the top quantile of PRS_E2_ (OR=0.95, p=0.51) (**Fig. 4f; Extended Data 13**). Similarly, the risk-increasing effect of a longer reproductive interval (>9 years) was observed in the low PRS_E2_ group (OR=1.26, p=1.3×10^-4^), but not in the high PRS_E2_ group (OR=1.02, p=0.73). Although the overall interaction term was not statistically significant, the protective impact of later age at menarche (>13 years) was limited to the low PRS_E2_ group (OR=0.85, p=3.9×10^-4^), but not among women with average PRS_E2_ (OR=0.96, p=0.098) or those in the top PRS_E2_ quantile (OR=0.95, p=0.27), suggesting that the protective influence of delayed menarche is attenuated by estrogen-modulating breast cancer risk variants. Despite the nominally significant interaction, the effect of parity was comparable in magnitude across PRS_E2_ strata.

## DISCUSSION

Here, we integrated functional genomics with epidemiological analyses to identify noncoding breast cancer risk variants whose regulatory activity is modulated by estrogen. Using MPRA, we identified 73 estrogen-modulated SNVs (emSNVs) showing significant genotype-by-estrogen interactions. Orthogonal validation through chromatin accessibility modeling, transcriptomics, and network analysis established that these variants operate through temporally regulated and mechanistically distinct pathways. Critically, emSNVs demonstrated population-level gene-environment interactions with reproductive risk factors in UK Biobank participants, where interactions were absent for matched non-estrogen-responsive variants. This demonstrates that functional profiling under disease-relevant conditions can identify variants with GxE interactions and point to their putative targets.

Most emSNVs (76%) showed peak activity at 6 hours post-estrogen treatment, mirroring ERα-mediated transcriptional kinetics. Chromatin accessibility at this timepoint showed strongest breast cancer heritability enrichment via S-LDSC, further supporting relevance.

Integration of MPRA with ChromBPNet revealed dual regulatory mechanisms: variants showing concordant effects (R=0.81) disrupt pioneer factor binding sites (FOXA, AP-1) that actively remodel chromatin, with MPRA-only variants affecting secondary factors (NF-κB, E2F) recruited to pre-accessible enhancers. This distinction may help explain assay-specific patterns and suggests that specific variants can modulate different regulatory layers, such as chromatin accessibility and TF recruitment. TF-MoDISco analysis revealed sequential TF programs across timepoints, suggesting estrogen triggers ordered waves of regulatory factor recruitment that emSNVs disrupt at specific steps. Protein interaction network analysis revealed emSNV target genes converge on histone variants (H4C1, H4C6, H4C11) modules including nucleosome assembly, mitochondrial metabolism, NF-κB signaling, and immune regulation, each of which are interconnected processes dysregulated in ER-positive and endocrine-resistant breast cancer. This convergence is consistent with estrogen inducing robust histone expression specifically in ER-positive cells. Thirty-eight identified network proteins are druggable targets, suggesting that while emSNVs cannot be readily modified, their downstream pathways represent potential intervention points for mitigating genetic risk in high-burden individuals.

The MPRA-informed PRS (PRS_E2_) showed comparable main effects to a control PRS of non-estrogen-responsive variants, yet only PRS_E2_ exhibited significant interactions with reproductive exposures. Risk effects of later age at first birth and longer reproductive interval were attenuated in women with high PRS_E2_, as were protective effects of later menarche. This pattern where hormonal risk factors primarily influence women with low genetic burden of estrogen-responsive variants suggests that constitutive genetic sensitivity to estrogen may buffer susceptibility to additional environmental modulation, potentially reflecting saturation effects where genetic predisposition dominates.

Prior GxE studies using PRS optimized for prediction of breast cancer risk identified interactions only with family history^2^, not hormonal exposures. Traditional PRS aggregate signals across multiple susceptibility pathways, making it challenging to isolate genetic effects operating through specific environmental mechanisms. Our approach differs by restricting the PRS to variants with functional response to the relevant hormonal exposure. Despite reducing the total number of variants, this approach improved the capacity to detect GxE by enriching for interaction effects among candidate SNVs selected based on marginal effects. The advantage of partitioning a standard PRS to increase power for GxE discovery is also supported by recent studies using pathway-specific PGS for liver biomarkers and adiposity^78,79^. Furthermore, compared to a PRS based on in-silico annotations or additional statistical associations, a functionally informed PRS improves power by reducing phenotypic heterogeneity and compensating for measurement error in self-reported exposures, providing a more precise and biologically interpretable. The absence of interactions for the control PRS comprised of variants from the same loci without estrogen-responsive function further demonstrates the specificity of the observed interactions to estrogen modulation rather than general regulatory activity.

Previous MPRA studies of breast cancer variants identified functional SNVs but did not test environmental modulation or temporal dynamics. Our work demonstrates that static assays may miss context-dependent effects critical for understanding mechanisms. Several emSNVs (rs9479091 in ESR1, rs11836367 affecting GATA3-NTN4) overlap known ER-bound enhancers, validating this approach while identifying novel estrogen-responsive elements. The distinction between accessibility-modulating versus recruitment-modulating SNVs, extended insight beyond variant prioritization to nominate regulatory principles active in hormone-responsive tissues.

The combination of MPRA, chromatin profiling, transcriptomics, and network analysis applied here provided convergent evidence for estrogen-responsive regulatory mechanisms. The observation that only 25% of emSNV target genes overlap with estrogen-responsive DEGs reflects the distinct biological processes captured by each approach: MPRA measures direct regulatory element activity, ChromBPNet models chromatin accessibility, and RNA-seq captures integrated steady-state expression influenced by multiple regulatory inputs. This partial overlap is expected given the layered nature of gene regulation and supports the premise that variants operate through both accessibility-dependent and accessibility-independent mechanisms. The use of MCF-7 cells proved helpful, given their well-characterized estrogen responsiveness and the fact that estrogen-responsive ATAC-seq peaks from these cells enriched for breast cancer heritability across all subtypes, including ER-negative and triple-negative disease, via S-LDSC.

This study has several limitations. Evaluation of PRSxE in the UK Biobank focused on women of European ancestry, the population with highest statistical power both for PRS and reproductive factor associations. While testing in additional populations would be valuable for assessing generalizability, the observed interactions are underpinned by mechanistic data and we expect that functional variants are likely to be conserved across ancestries, allowing extension to diverse cohorts, although differences in LD and allele frequencies may necessitate direct validation.

This study establishes that systematic functional profiling under disease-relevant environmental conditions identifies variants with population-level gene-environment interactions. The convergence of molecular and epidemiological evidence demonstrates that in vitro GxE predicts in vivo GxE when variants are selected based on functional response. This framework may be applied to other diseases and modifiable risk factors by profiling variants with differential response to dietary, inflammatory, or other stimuli to identify gene-environment interactions that may be currently obscured in standard SNVxE and PRSxE analyses.

Elucidating pathways through which genetic and environmental factors interact enable risk stratification and identifies potential intervention targets. Women with high PRS_E2_ may benefit from intensified screening regardless of traditional risk factors, while those with low PRS_E2_ and high-risk exposures represent a distinct vulnerable group. This framework provides an approach to investigate gene-environment interactions in polygenic diseases and may help inform future strategies for precision prevention.

## METHODS

### Genetic variants associated with breast cancer

Lead SNVs from the GWAS catalog^16^ (v.3.0) were collected by filtering for breast cancer terms (Supplemental Table 1) and selecting for those at genome-wide significance level 5e-08. All SNVs in LD with the lead SNVs (r2 > 0.8) were identified with LDlink within the populations “EUR”, “AFR”, “AMR”, “EAS”, “SAS” and “ALL” separately. This expanded the 583 lead SNVs to 19,426 SNVs. Without considering indel mutations, this represented 257 LD-independent or clumped SNVs (at LD r2 < 0.10). SNVs were filtered for putative regulatory activity if they overlapped a union set of peaks from DNAse-seq (ENCFF594NFE), ATAC-seq (ENCFF749WUW, ENCFF402SHW), H3K4me1 ChIP (ENCFF245WLI, ENCFF449OJY), H3K27ac ChIP (ENCFF804TDD, ENCFF481ARW) in female breast epithelium tissue. Seven additional SNVs (rs10069690, rs12998806, rs13074711, rs17530068, rs2284378, rs8100241, and rs9383938) and their linked SNVs were included from literature. This resulted in a final set of 1604 SNVs or 73 LD-independent regions (at an r2 < 0.10). SNVs were filtered if they contain an indel greater than 3 bp. The final library was built by including 162 bp of genomic sequence centered on the SNV. Lastly, if multiple SNVs were within 81 bps, sequences representing the most common haplotypes according to the 1000 Genomes project were included in the library but were not analyzed for this study. Each genomic and random fragment was barcoded with 10 random 16bp barcodes. This library was prepared as previously described^80^ and contained 37,080 oligos and included 1,604 SNVs.

### Lentivirus production

Lenti-X 293T cells (Takara) were maintained in DMEM (10% FBS and 1% penicillin– streptomycin at 37 °C with 5% CO2). For virus preparation, 293T cells were seeded at 12 million cells per 15cm plate and the following day transfected with 18.75ug pCMV-dR8.91, 6.25ug pMD2.G, and 25ug of the MPRA library using OptiMEM (Thermo Fisher) and Lipofectamine 3000 (Invitrogen). The supernatant was collected 48 hours later, filtered with a 0.44um filter, and concentrated 50x using LentiX Concentrator (Takara).

### Massively Parallel Reporter Assays (MPRA)

To assess how estrogen influences the transcriptional activity of BRCA risk-associated SNVs, MPRA was performed in the presence and absence of estrogen. A barcoded MPRA library^13^ using SNV-centered 150bp native genomic fragments was first cloned into the pGreenFire lentivector (**Fig. 1A**). The resulting library preserved input sequence complexity, displaying 99.6.% allelic coverage and 21.53 skew (**Supp Fig. 1A**). Lentiviral transduction was then performed into an ER+ BRCA cell line MCF-7, as primary human mammary epithelial cells (HMECs) do not show robust ER expression^81,82^, then cells were grown in the presence and absence of estrogen (**Fig. 1A**) prior to harvesting and RNA-sequencing. Analysis of the resulting data was performed to identify breast cancer risk SNVs modulated by estrogen. SNVs not in dbSNP were excluded as candidates for further exploration. Oligo libraries were synthesized by Agilent, PCR amplified and ligated into pGreenFire-mCMV (EF1a-puro, System Biosciences) as described previously.^13,80^ For each MPRA biological replicate, 4-5 million MCF7 were transduced in 15 cm plates in DMEM (10% FBS and 1% penicillin–streptomycin at 37 °C with 5% CO2) with the addition of 5 µg/mL polybrene. Cells were selected in 2.0 µg/mL puromycin for 24-48 hours after transduction. Once selected, cells were seeded for 10 nM estrogen (Sigma, estrogen758) treatment for 6 and 24 hours, representing concentration and time points commonly used in MCF7 in-vitro studies for transcriptomic profiling.^83^ For sequencing library construction, total RNA was isolated using Qiagen’s RNeasy Plus kit (Qiagen, #74136) and mRNA subsequently purified using Dynabeads mRNA DIRECT purification kit (ThermoFisher, 61011). Reverse transcription was performed with 500 ng mRNA, 100 nM primer and SuperScript IV (ThermoFisher, 18090050) according to the manufacturer’s protocol. Reactions were incubated with 1 µL Themolabile Exonuclease I (NEB, M0568L) at 10 min at 37°C, followed by inactivation at 85° for 5 min and cDNA purification with AMPure XP beads (Beckman Coulter, A63880) at a 1:1.1 sample-to-bead ratio. PCR amplification was performed in 50 µL reactions containing 5 µL cDNA, SYBR green (ThermoFisher, S7563), and PrimeStar Max DNA Polymerase (Takara, R045B). Reactions stopped in the early exponential phase, pooled, concentrated, gel purified and sequenced. Sequencing was performed on a NovaSeq 6000, paired-end 150 bp reads, with an average of 14 million reads per sample.

*MPRA analysis*. MPRA libraries were designed, synthesized, and assayed as previously described.^13,80,84^ UMIs and barcodes were extracted from sequencing reads using UMI-tools12 v. 1.1.5. Bowtie13 v. 1.3.1 was used to map barcodes to a reference index of the barcode library allowing for up to one mismatch. The number of UMIs per guide was calculated from the mapped read files using the UMI-tools count program with the “directional” method. MPRAnalyze14 v. 1.9.1 was used to determine differentially active genomic fragments. SNVs were only analyzed if they had at least 5 barcodes detected in the plasmid DNA library for both the alt and ref allele. Differential activity for the individual timepoints was calculated using “analyzeComparative()” with DNA design “∼barcode”, RNA full design “∼allele” and reduced design “∼1”. For the analysis of estrogen-allele interactions, the RNA full design was “RNA counts∼ estrogen condition + allele + estrogen condition:allele” and reduced design “ RNA counts∼ estrogen condition + allele”.

### Assignment of Putative Target Genes to SNVs

To nominate potential emSNV-modulated targets, putative target genes (**<u>ptGenes</u>**) were assigned to emSNVs. Nearest gene was assigned to genes within 100Kb of each SNV. Proximity-based ptGenes were defined as those with promoters located within 5 kb of an emSNV. Second, expression quantitative trait loci (**<u>eQTL</u>**) data were incorporated from GTEx (v8) for cell types relevant to the cancer associated with each emSNV and from eQTLGen (version 2019-12-11)^85^ and pancanQTL. HiChIP data for H3K27ac, an active enhancer mark, was used to identify promoter-emSNV interactions. Promoters were defined as regions within 5 kb of annotated transcription start sites, and HiChIP anchors were annotated with genes whose promoters overlapped these regions. ptGenes were assigned to specific emSNVs based on interactions where the emSNV and promoter were located at opposite ends of a chromatin loop. These interactions were filtered to include only those occurring in cell types relevant to the disease associated with the emSNV as well as target mRNAs modulated by estrogen identified by RNA-seq above. This integrative approach nominated emSNV ptGenes (**Extended Data 3).**

### Training ChromBPNet

We used the Encode ATAC-seq pipeline to process ATAC-seq data including aligning reads and calling peaks. The pipeline is documented on github: https://github.com/ENCODE-DCC/atac-seq-pipeline. The pipeline was run with default settings. For each sample we trained a ChromBPNet model using the steps described in the official ChromBPNet repo: https://github.com/kundajelab/chrombpnet. In summary, we used ‘chrombpnet prep nonpeaks’ to generate GC-matched negative regions for each sample. We then trained a bias model using ‘chrombpnet bias pipeline’ with default settings. Lastly, we trained an accessibility model using ‘chrombpnet pipeline’. All downstream analyses were performed on the accessibility models.

### De novo motif discovery

In order to identify motifs learned by ChromBPNet we used tfmodisco-lite v2.4.0 (https://github.com/jmschrei/tfmodisco-lite). For each model, base-pair contribution scores were calculated using the ‘deep_lift_shap function’ in tangermeme v0.5.0 in peak regions (https://github.com/jmschrei/tangermeme). We then passed these contribution scores into tfmodisco-lite along with one-hot encodings of the sequence regions and ran the package as described in the repo with default parameters. We also used tfmodisco-lite to identify motifs existing near variants of interest. For regions of interest, we repeated the same process as above but ran tfmodisco with a window size (-w) of 100 base pairs.

### Variant scoring

To quantify the regulatory impact of non-coding variants, we used bias-corrected ChromBPNet model trained to predict chromatin accessibility from DNA sequence. For each variant, we extracted a 2,145 bp window centered on the variant from the reference genome and generated predictions for both reference and alternate alleles.

We computed several metrics to quantify the effect of each variant, including the log fold change (logFC) in predicted accessibility, the Jensen–Shannon divergence (JSD) between predicted accessibility profiles, and their product (logFC × JSD). These were calculated from both the total predicted accessibility (counts) and the base-resolution accessibility profiles.

To estimate empirical p-values, we also computed these scores for a set of randomly shuffled variants matched to the input variants. Variants with insertions or deletions were handled by adjusting JSD scores to account for sequence length differences.

Final outputs included per-variant scores and associated p-values, stored in a tab-delimited file. We used default parameters for all options.

### MPRA-informed polygenic risk scores

To derive an MRPA-informed PRS that is more likely to capture GxE effects, linkage disequilibrium (LD) clumping was performed on 73 emSNVs to select independent SNVs with an LD threshold (r²) of 0.1 and a 1 Mb window using a subset of 10,000 European ancestry UK Biobank participants as the reference panel. PRS_E2_ was calculated as the weighted sum of risk-increasing alleles with weights corresponding to the marginal log odds ratio (OR) for breast cancer obtained from Zhang et al. GWAS meta-analysis based on 133,384 breast cancer cases and 113,789 controls from the Breast Cancer Association Consortium (BCAC).

As a comparison and negative control, we derived PRS_E2-NEG_ comprised of SNVs that did not show evidence of response to estrogen stimulation in the MPRA analysis. Specifically, we filtered SNVs that had a p-value > 0.8 in the 6-hour interaction with estrogen from the MPRA panel and selected one SNV for each variant in the PRS_E2_ from those SNVs on the same chromosome at least 50 Mb away. The effect sizes for these SNVs were again obtained from the meta-analyzed risk allele weights from the BCAC summary statistics.

### Gene-Environment Interaction Analysis in the UK Biobank

Interaction analyses were performed in the UK Biobank (UKB), a population-based prospective cohort of individuals aged 40–69 years, enrolled between 2006 and 2010. ^86^ Health outcomes were ascertained through linkages to national cancer registries, hospital in-patient encounters, and national mortality records. Details of the quality control and phenotyping process have been previously described.^87,88^ Briefly, analyses were restricted to individuals with concordant self-reported gender and genetically inferred sex. Participants were determined to be of European ancestry based on self-reported ethnicity and genetic ancestry PCs. Individuals with values for PC1 or PC2 that were outside of 5 standard deviations from the population mean were removed. Relatedness was inferred using KING^89^ and one individual from each pair of first-degree relatives was removed, preferentially retaining cancer cases. Individuals with at least one recorded diagnosis of a borderline, in situ, or malignant primary cancer based on International Classification of Diseases (ICD)-9 or ICD-10 codes were defined as cases.^87,88^Follow-up time in the current analysis extends to September 2020. Analyses were limited to participants free of breast cancer and post-menopausal at baseline. The final cohort consisted of 13, 026 incident breast cancer cases and 108, 265 controls with genetic, reproductive, and phenotypic data.

Risk factors identified within the cohort include use of hormone replacement therapy (ever/never), oral contraceptive use (ever/never and duration), age at menarche, age at menopause, nulliparity, and age at first birth. Two additional exposure variables were generated that combined consequential, estrogen exposure-related milestones: time from menarche to first birth and time from menarche to menopause. Continuous exposure variables were dichotomized using cutoffs derived from the literature:^90^ age at menarche at 13 years, age at first birth at 30 years, and the time from menarche to first birth at 9 years. Associations for established breast cancer risk factors were estimated using logistic regression models adjusted for age at baseline.

PRS_E2_ was analyzed as both a standardized continuous and categorical variable. For categorical analyses, the PRS_MRPA_ was categorized into Low (≤20th percentile), Average (20th– 80th percentile), and High (≥ 80^th^ percentile), with the Average PRS_E2_ group serving as the reference category. Logistic regression models were fitted to examine the effects of the interactions between PRS_MRPA_ and each of the reproductive or hormonal risk factors on breast cancer risk **(Supplementary Table X**). Interaction testing was only performed for risk factors with statistically significant main effects in UKB. All models that included PRS_E2_ or PRS_E2-NEG_ were adjusted for age at baseline and the first 10 genetic ancestry principal components (PCs). Statistically significant interactions were determined based on the Wald statistic for the (PRS × risk factor) interaction term, which tests the null hypothesis that the corresponding regression coefficient is zero. The overall statistical significance of interaction terms that included the categorized PRS was determined using a chi-squared test. All statistical analyses were performed using R v.4.2.2.

### Gibson cloning for emSNVs

emSNVs were ordered as IDT ultramers. IDT Ultramers (4 nmol) were resuspended in 40 μl of nuclease-free water to achieve a final concentration of 100 μM. The stock solution was diluted 1:50 by mixing 2 μl of the resuspended ultramer with 98 μl of 1X NEBuffer 2, resulting in a 2 μM working solution. Single-stranded DNA (ssDNA) oligos were further diluted to a final concentration of 0.2 μM in 1X NEBuffer 2. For DNA assembly, a 10 μl reaction was prepared by combining 5 μl of ssDNA oligo (0.2 μM), 30 ng of restriction enzyme-linearized vector, and ddH2O, followed by the addition of 10 μl of NEBuilder HiFi DNA Assembly Master Mix (New England Biolabs). The reaction was incubated at 50°C for 1 hour. After assembly, 2 μl of the reaction product was transformed into NEB 10-beta chemically competent *E. coli* cells according to the manufacturer’s protocol. Transformed cells were plated on LB agar containing ampicillin and incubated overnight at 37°C. The following day, 10 colonies were picked and grown in LB medium containing ampicillin. Plasmid DNA was purified from these cultures using a standard miniprep protocol and sequenced to verify successful assembly.

### RNA-seq Analyses

Total RNA was extracted and sent to Novogene for mRNA sequencing using their mRNA-Sequencing service. Library preparation was performed using poly(A) selection and a non-directional protocol to enrich for mRNA transcripts. Sequencing was conducted on the NovaSeq X Plus platform, generating 30 million paired-end reads per sample with an average output of 9 Gb per sample. Quality control of raw sequencing data and subsequent data analysis were performed following the workflow described in the GitHub repository rna-seq-star-deseq2. Briefly, reads were aligned to the reference genome using STAR, and differential gene expression analysis was performed using DESeq2. All analyses were conducted with default parameters unless otherwise specified, ensuring reproducibility and consistency with established RNA-seq pipelines.

### Luciferase Assays

MCF7 cells were cultured in DMEM without phenol red supplemented with charcoal-stripped fetal bovine serum (CS-FBS; Sigma) for 48 hours prior to transfection to reduce endogenous estrogen levels. On the day before transfection, cells were seeded in 24-well plates at 60–70% confluency. For each SNP, six tubes were prepared for reference and alternate alleles, along with tubes containing an empty vector as a control. Each tube contained 125 μl of OptiMEM, 5 μl of pGL3-Rluc plasmid (5 ng/μl stock), and 3.6 μl of pGL4-Fluc plasmid (350 ng/μl stock) or the empty vector control. To each tube, 5 μl of FuGENE-4K was added directly into the OptiMEM and immediately mixed by gentle tapping to ensure homogeneity while avoiding contact of FuGENE with the tube walls. The reaction was incubated at room temperature for 15 minutes before 50 μl of the transfection mix was added to duplicate wells for each condition.

The following day, cells were treated with 10 nM estradiol (estrogen; Sigma) or vehicle control (ethanol) for 24 hours, resulting in four conditions per SNP: reference (no estrogen), alternate (no estrogen), reference (+estrogen), and alternate (+estrogen). Wells transfected with the empty vector were included as controls for normalization. After treatment, cells were lysed in 150 μl of 1X Passive Lysis Buffer (PLB; Promega), and 10 μl of lysate from each sample was used for a dual luciferase reporter assay. The dual luciferase assay was performed using the Promega Dual-Luciferase® Reporter Assay System according to the manufacturer’s instructions. Firefly luciferase activity (pGL4-Fluc) was measured as the primary reporter, followed by the quantification of Renilla luciferase activity (pGL3-Rluc) as an internal control. Relative luciferase activity was calculated by first normalizing firefly luciferase activity to Renilla luciferase activity for each sample. These values were further normalized to the empty vector control to account for baseline luminescence and subtracting blank well readings to correct for background signal. Measurements were performed in a luminometer, and each condition was tested in biological duplicates to ensure reproducibility.

### ATAC-seq peak processing and generation of LDSC annotations

Raw ATAC-seq data from individual technical replicates were processed independently. For each replicate, tagAlign.gz files were obtained from the ENCODE-style pipeline output and converted to BAM format. Each BAM file was indexed using samtools index. To define a unified set of peaks across all conditions, we collected the narrowPeak files for each replicate, corresponding to the peak calls with p < 0.01. Peak regions were decompressed, concatenated, and then merged to produce a non-redundant global consensus peak set. Read counts overlapping these consensus peaks were quantified using bedtools multicov, with each BAM file from each technical replicate treated independently. This produced a matrix of read counts across ∼330,000 consensus peaks for all replicates and conditions. This matrix was subsequently used as input for differential accessibility analysis using the edgeR package, allowing for statistical modeling of dispersion across replicates. We obtained high-confidence ATAC-seq peaks from control (C1), estrogen 6-hour (s6R1), and 24-hour (s24R1) conditions using irreproducible discovery rate (IDR) thresholding across replicates. To generate LDSC annotations, we retained the genomic coordinates (chromosome, start, end) for high-confidence ATAC-seq peaks from each narrowPeak file.

### Stratified LD score regression

From subtype-specific GWAS of breast cancer in the European population (ref 2017, 2020 papers), we focused on subtypes with SNP heritability > 0.04; ER-positive, ER-negative, TNBC, luminal A, liminal B, luminal B Her2 negative breast cancer, together with overall breast cancer. (add citations to 2017 or 2020 papers for each subtype). To estimate SNP heritability, we utilized effective sample size of each subgroup in GWAS reported in the original paper.

We performed ATAC-seq in three different conditions in MCF7; without stimulation, 6 hours after estrogen stimulation, and 24 hours after stimulation. For each condition, we called reproducible narrow peaks following ENCODE pipeline^91^.Further, we defined peaks gained after estrogen stimulation using bedtools subtract module with -f 0.95 option. Namely, we defined peaks for which more than 5% emerged after stimulation as gained peaks. We calculated the LD scores of the SNPs overlapping with the peaks using the 1000G Phase 3 population reference.

We added each annotation to the baseline1.1 model and regressed against trait chi-squared statistics using HapMap3 SNPs with the stratified LD score regression package v.1.0.1 to estimate the heritability enrichment.

## Data Availability

All data produced in the present study are available upon reasonable request to the authors.
GEO data: GSE329425, GSE329426, and GSE329424.

## Data availability

Raw and processed MPRA sequencing data, RNA-Seq, and ATAC-seq data from this study are available through the Gene Expression Omnibus under accession number GSE329425, GSE329426, and GSE329424 respectively.

## Code availability

Code related to the MPRA analyses in this study can be found on GitHub at https://github.com/IbbyElfaki/mpra-gwas-builder

## Acknowledgments

We thank G. Rayant and K. Fields for their generous support and helpful discussions. This work was supported by USVA Office of Research and Development BX001409 (P.A.K.) and by NIAMS/NIH AR076965 and AR43799 to PAK,

## Author contributions

I.E, L.K, and P.A.K. conceptualized the project. I.E, D.L.R, L.N.K. R.M.M contributed to the design of the MPRA. I.E performed MPRA experiments. I.E, S.M X.Y, D.L.R performed validation and follow-up experiments. I.E, L.D, M.F, K.O, M.O, D.F.P, S.S, T.N analyzed the data. I.E L.N.K,K.W, N.S.A, J.M.M, T.F, R.M.M, L.K, and P.A.K. guided methodology development, experiments and data analysis. I.E, L.K, and P.A.K. wrote the manuscript with input from all authors

## Competing interests

Authors declare that they have no competing interests.

## Supplementary Materials

Extended Data:

1. MPRA statistics
2. MotifbreakR data
3. SNP-putative gene
4. RNAseq data
5. LDSC statistics
6. ChromBPNet control SNP scores
7. ChromBPNet 6hr SNP scores
8. ChromBPNet 24hr SNP scores
9. Baseline characteristics for included participants in UK Biobank
10. Breast Cancer summary statistics across breast cancer subtypes
11. MPRA-informed polygenic risk score associations in the UK Biobank.
12. Interactions between estrogen-related breast cancer risk factors and continuous PRS-estrogen observed in post-menopausal women in the UK Biobank.
13. Interactions between estrogen-related breast cancer risk factors and discretized PRS-estrogen observed in post-menopausal women in the UK Biobank.
14. Interactions between estrogen-related breast cancer risk factors and continuous PRS-estrogen-NEG observed in post-menopausal women in the UK Biobank.
15. Interactions between estrogen-related breast cancer risk factors and discretized PRS-estrogen-NEG observed in post-menopausal women in the UK Biobank.

## Notes

### Competing Interest Statement

The authors have declared no competing interest.

